# KNOWLEDGE OF METABOLIC SYNDROME, LIFESTYLE RISK BEHAVIOURS AND PREVENTIVE PRACTICES AMONG WOMEN ATTENDING KASOA POLYCLINIC, GHANA: A CROSS-SECTIONAL STUDY

**DOI:** 10.64898/2026.10.14.26365545

**Authors:** Grace De-Graft Aitee, Evelyn Owusu, Abena Ntiriwaa Cann, Edmund Teye Borkor, Abubakar Iddrisu Siddiq, Afua Pokuaa Kwakye, Ernest Alfred Kwaku Ampong

**Affiliations:** Central Regional Health Directorate, Ghana Health Service; College of Nursing and Midwifery, Koforidua, Ministry of Health; Northern Regional Health Directorate, Ghana Health Service; Department of Clinical Optometry, University of Cape Coast; Department of Public Health, Faculty of Medical Sciences, Presbyterian University, Ghana

**Keywords:** metabolic syndrome, women, knowledge, lifestyle, hypertension, diabetes, Ghana

## Abstract

**Background:** Metabolic syndrome is a cluster of cardiometabolic risk factors that may increase the risk of cardiovascular disease and type 2 diabetes, and African studies report a higher prevalence in women than in men. Prevention may depend, in part, on whether women recognise the condition and its modifiable risk factors. Relatively little appears to be known about this among women using primary-level health facilities in fast-growing peri-urban areas of Ghana.

**Objective:** To assess knowledge of metabolic syndrome, describe lifestyle risk behaviours, self-reported cardiometabolic conditions and preventive practices, and examine selected lifestyle correlates of self-reported diabetes and hypertension among women attending Kasoa Polyclinic, Ghana.

**Methods:** A descriptive cross-sectional survey was conducted in June and July 2024 among 174 women aged 18–45 years recruited by convenience sampling. A structured, interviewer-assisted questionnaire covered knowledge, lifestyle behaviours, self-reported diagnoses and preventive practices. Data were summarised as frequencies and percentages, and associations were examined with Pearson’s chi-square test.

**Results:** Only 37 women (21.3%) described metabolic syndrome as a condition that raises the risk of heart disease and type 2 diabetes, 40 (23.0%) identified central (apple-shaped) fat as the high-risk pattern and 31 (17.8%) knew that dietary recommendations exist. Most reported no weekly physical activity (116; 66.7%), daily sugary drinks (109; 62.6%), daily processed or fast food (100; 57.5%), high stress (103; 59.2%), and less than six hours’ sleep (94; 54.0%). Self-reported diagnoses were hypertension in 60 (34.5%) and type 2 diabetes in 49 (28.2%). Fifty-nine (33.9%) had never received advice on prevention and 105 (60.3%) did not monitor their weight or waist. Self-reported diabetes was associated with short sleep (36.2% among those sleeping under six hours; p = 0.035) and with stress level (p = 0.028), being most frequent among women reporting low stress. Self-reported hypertension was more common with high stress (42.7%; p = 0.022) and with some physical activity (p = 0.005).

**Conclusion:** Women attending this polyclinic appeared to have limited understanding of metabolic syndrome and a high burden of modifiable risk behaviours. Routine counselling on diet, activity, sleep, and weight monitoring at the polyclinic may be warranted. The bivariate, self-reported associations should be interpreted cautiously.

## 1. Introduction

Metabolic syndrome describes the co-occurrence in one person of central obesity, raised blood pressure, raised fasting glucose, raised triglycerides, and low high-density lipoprotein cholesterol (1). Under the harmonised definition, any three of these five components establish the diagnosis (1). The syndrome may be clinically important because it identifies people at increased risk of cardiovascular disease and type 2 diabetes, and its components appear to be largely driven by diet, physical inactivity, and excess weight (2).

The condition is common worldwide (3). A meta-analysis of data from more than 28 million adults estimated a global prevalence of 12.5% to 31.4%, depending on the definition used (3). In Africa, a pooled analysis estimated a prevalence of 32.4%, with a higher estimate in women (36.9%) than in men (26.7%) and a prevalence of 33.1% in West Africa (4). A systematic review of Ghanaian studies likewise found metabolic syndrome to be frequent among adults considered apparently healthy (5). Its components are also common in Ghana, where hypertension is widespread among adults (6) and diabetes has become an established public health concern (7).

Several lifestyle behaviours are associated with metabolic syndrome and its components. Regular intake of sugar-sweetened beverages is associated with a higher risk of both metabolic syndrome and type 2 diabetes (8). Short sleep is associated with a higher incidence of type 2 diabetes (9), and psychosocial stress with hypertension (10). The World Health Organization recommends that adults undertake at least 150–300 minutes of moderate-intensity aerobic activity each week and limit sedentary time (11). Rapid urbanisation in Ghana may have made processed foods, sugary drinks and sedentary work more common, particularly in peri-urban towns (12).

Prevention requires that women understand what metabolic syndrome is, which behaviours raise their risk and what they can do about it. The Health Belief Model suggests that people are more likely to act on a health threat when they perceive themselves to be susceptible, regard the condition as serious and believe that action will help (13). Knowledge of the condition and its risk factors may constitute a precondition for these perceptions. Most Ghanaian research on metabolic syndrome has measured its prevalence (14), and less is known about what women attending primary-level facilities know about it or how they live in relation to its risk factors.

Kasoa, the capital of the Awutu Senya East Municipality in the Central Region, is one of the fastest-growing urban areas in Ghana, with overcrowding and pressure on services (15). Women attending its polyclinic come from a population undergoing the rapid lifestyle change that favours metabolic risk. Published data on knowledge of metabolic syndrome or related behaviours among women in this setting appear to be lacking.

This study therefore assessed knowledge of metabolic syndrome among women aged 18–45 years attending Kasoa Polyclinic. It also described their lifestyle risk behaviours, self-reported cardiometabolic diagnoses and preventive practices, and examined whether selected lifestyle factors were associated with self-reported type 2 diabetes and hypertension.

## 2. Methods

### 2.1 Study design and setting

A descriptive cross-sectional survey was conducted at Kasoa Polyclinic in the Awutu Senya East Municipality, Central Region, Ghana. The polyclinic is a 45-bed facility with medical, surgical, maternity, antenatal, dental, eye, emergency, laboratory, and pharmacy services, and serves the rapidly growing Kasoa area. Data were collected over 15 days in June and July 2024. The report follows the STROBE statement for cross-sectional studies (16).

### 2.2 Study population

Women aged 18–45 years who attended the polyclinic for any service, including routine check-ups and consultations, and who were able to understand the study and consent were eligible. Women employed as health professionals at the polyclinic were excluded because their training could have influenced their knowledge, as were women outside the age range and those unable to give informed consent.

### 2.3 Sample size and sampling

The sample size was calculated with Cochran’s formula, n = z²pq/d² (17), using z = 1.96 for 95% confidence, a margin of error (d) of 0.05 and an assumed proportion (p) of 0.13. The proportion was taken from a previous knowledge survey on diabetes (18), because no estimate of metabolic syndrome knowledge among Ghanaian women was available. The calculation yielded 173.8, which was rounded to 174. Recruitment proceeded by convenience sampling in waiting areas or after consultations, continuing until 174 women had been recruited. Data collection was facilitated through a structured questionnaire administered by trained research assistants to ensure consistency in capturing demographic profiles and self-reported health statuses. The survey instrument further incorporated sections on dietary habits and physical activity levels to identify specific modifiable lifestyle factors, utilizing validated food frequency questionnaires similar to those employed in other regional studies of cardiometabolic risk (19,20).

### 2.4 Data collection

A structured questionnaire was developed from the study objectives and the literature and reviewed by the academic supervisor for content validity. It was pre-tested among 30 women at the Mother and Child Hospital and revised before use. The questionnaire had four sections: socio-demographic characteristics; knowledge of metabolic syndrome, its risk factors and complications; lifestyle factors and previous diagnoses; and preventive practices. Responses were entered into an online form. Five research assistants with bachelor’s degrees in nursing or public health, trained for 16 hours over two days, administered the questionnaire. For women who could not read English, research assistants read and explained the questions in Twi and recorded their responses.

### 2.5 Study variables and operational definitions

#### Knowledge

Knowledge was assessed with single items and reported item by item. A response was regarded as correct when it agreed with current understanding of metabolic syndrome: describing it as increasing the risk of heart disease and type 2 diabetes; identifying central (apple-shaped) fat distribution as the higher-risk pattern; recognising a role for genetic factors; recognising each of five lifestyle factors (sedentary lifestyle, poor diet, excessive alcohol, smoking, and inadequate sleep) and five conditions (obesity, high blood pressure, high cholesterol, diabetes, and polycystic ovary syndrome) as risk factors; recognising five complications (cardiovascular disease, type 2 diabetes, stroke, kidney disease, and certain cancers); agreeing that weight loss reduces risk; and knowing that dietary recommendations exist. The question on the age group most susceptible was excluded from scoring because the risk increases with age (21) and no single option is correct.

#### Lifestyle factors

These were weekly frequency of physical activity, sedentary work, frequency of sugary drinks and of processed or fast foods, self-rated stress on a three-point scale (low, moderate, high), usual way of coping with stress, smoking, and usual hours of sleep per night.

#### Self-reported conditions

Women were asked whether they had ever been diagnosed with type 2 diabetes, high blood pressure, high cholesterol, or obesity. No clinical, anthropometric, or laboratory measurements were taken, and metabolic syndrome itself was not diagnosed.

#### Preventive practices

These were the recommended frequency of exercise for people with metabolic syndrome, receipt of advice from a health professional, following a balanced diet, stress-reducing activities, prescribed medication or supplements, frequency of blood pressure checks and monitoring of waist circumference and body weight.

### 2.6 Data management and statistical analysis

Completed questionnaires were checked daily for completeness and consistency. Data were analysed in Statistical Package for the Social Sciences (SPSS) version 26. Age is reported as mean and standard deviation, and categorical variables as frequencies and percentages of all 174 respondents unless otherwise stated. Associations between four lifestyle factors (physical activity, stress, sleep and processed or fast food) and self-reported type 2 diabetes and hypertension were examined with Pearson’s chi-square test. For this manuscript, the chi-square statistics were recomputed from the reported cross-tabulations to confirm them, and the proportion of women with each condition was calculated within each category of the lifestyle factor. All expected cell counts were five or more. Statistical significance was set at p < 0.05. Multivariable analysis was not performed, so the associations are unadjusted.

### 2.7 Ethical considerations

The study was approved by the Ghana Health Service Ethics Review Committee (GHS-ERC: 047/04/24; approved 25 June 2024). An introductory letter was obtained from the Ghana College of Nurses and Midwives, and permission to use the facility was granted by the management of Kasoa Polyclinic and the Awutu Senya East Municipal Health Directorate. Written informed consent, by signature or thumbprint, was obtained from every participant after the study had been explained in English or Twi. Participation was voluntary; no names were recorded and participants could withdraw at any time without consequence.

## 3. Results

### 3.1 Participant characteristics

All 174 women were aged 18–45 years, with a mean age of 35 years (standard deviation [SD], 7); half (87; 50.0%) were aged 30–39 years. Ninety-two (52.9%) had tertiary education and 25 (14.4%) had no formal education. Of the 173 who reported their residence, 111 (64.2%) lived within Kasoa. Professionals (46; 26.4%) and traders (41; 23.6%) were the largest occupational groups, and 26 (14.9%) were unemployed (Table 1).

**Table 1.** Socio-demographic characteristics of participating women, Kasoa Polyclinic, 2024 (N = 174)

| Characteristic | Category | n (%) |
| --- | --- | --- |
| Age group (years) | 18–19 | 3 (1.7) |
|  | 20–29 | 39 (22.4) |
|  | 30–39 | 87 (50.0) |
|  | 40–45 | 45 (25.9) |
|  | Mean (SD) | 35 (7) |
| Education | No formal education | 25 (14.4) |
|  | Basic | 24 (13.8) |
|  | Senior high school | 33 (19.0) |
|  | Tertiary | 92 (52.9) |
| Residence (n = 173) | Within Kasoa | 111 (64.2) |
|  | Outside Kasoa | 62 (35.8) |
| Occupation | Professional | 46 (26.4) |
|  | Trader | 41 (23.6) |
|  | Homemaker | 29 (16.7) |
|  | Unemployed | 26 (14.9) |
|  | Vocational | 23 (13.2) |
|  | Farmer | 9 (5.2) |

SD, standard deviation. <sup>a</sup> One respondent did not report residence. Percentages may not sum to 100 because of rounding.

### 3.2 Knowledge of metabolic syndrome

Knowledge of what metabolic syndrome is appeared limited. The most common description (66; 37.9%) was that it concerns rapid weight loss through food restriction, and only 37 women (21.3%) described it as a condition that increases the risk of heart disease and type 2 diabetes. Forty (23.0%) identified central (apple-shaped) fat as the higher-risk pattern, while 69 (39.7%) chose pear-shaped fat. A role for genetic factors was recognised by 56 (32.2%), and 72 (41.4%) regarded women aged 50 years and above as most susceptible (Table 2).

**Table 2.**
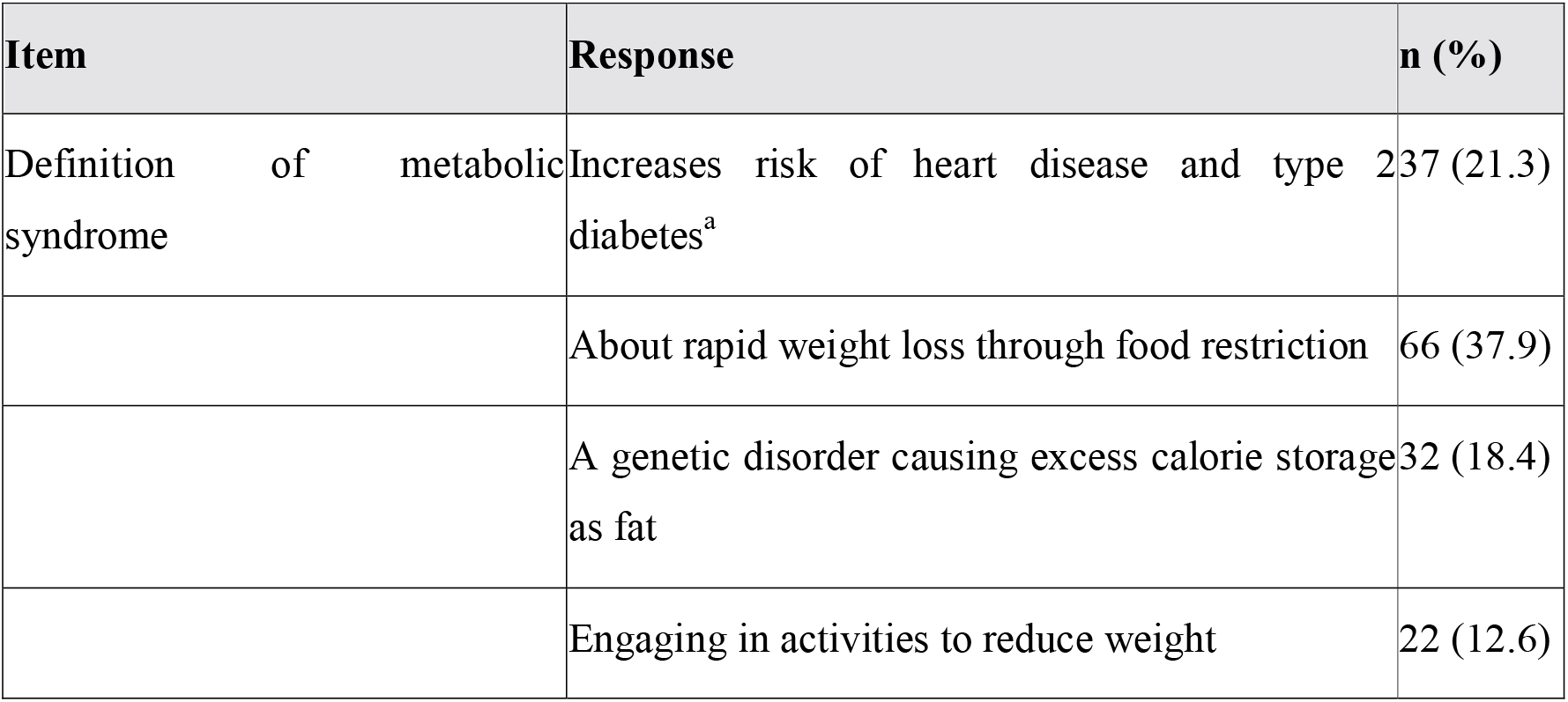
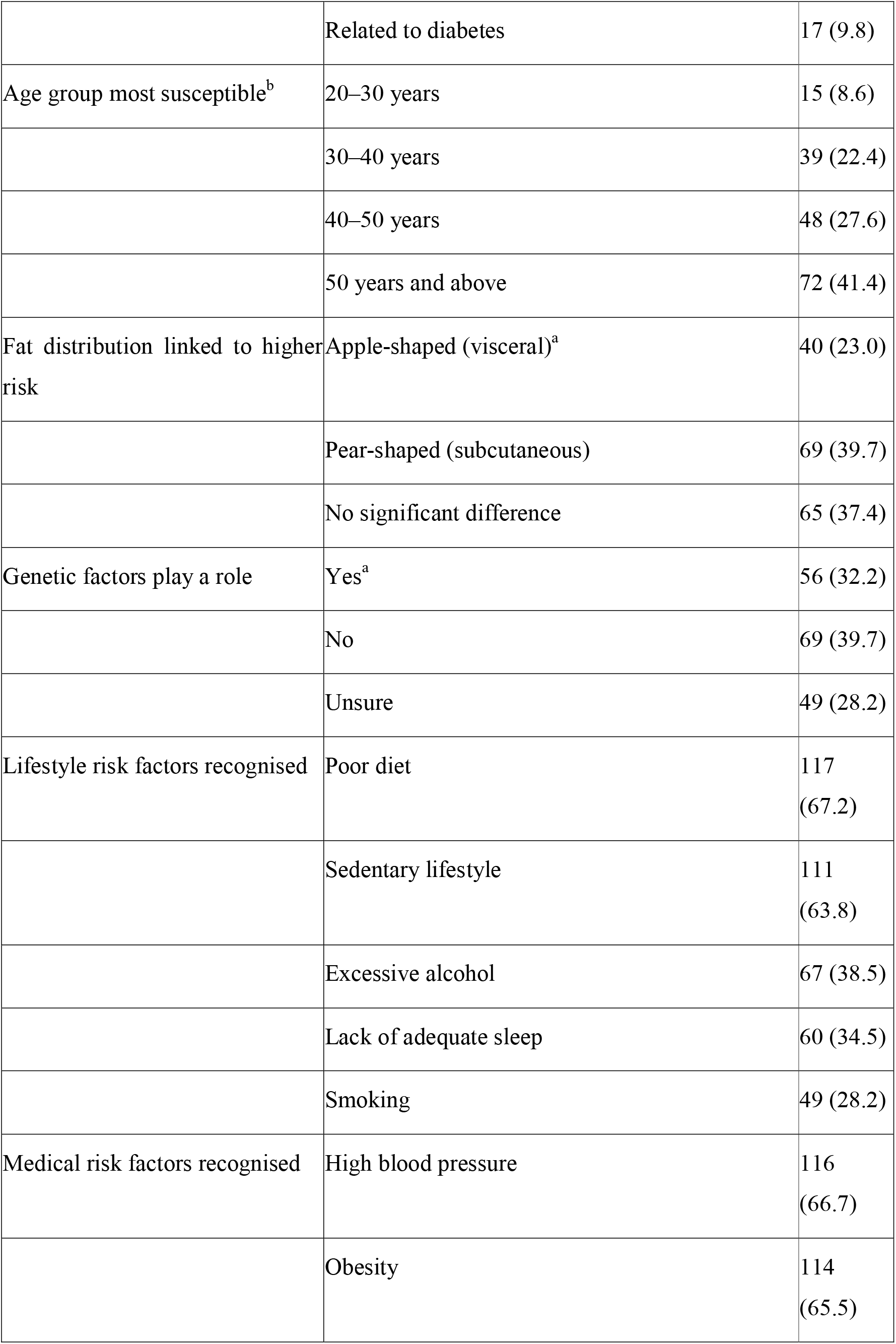
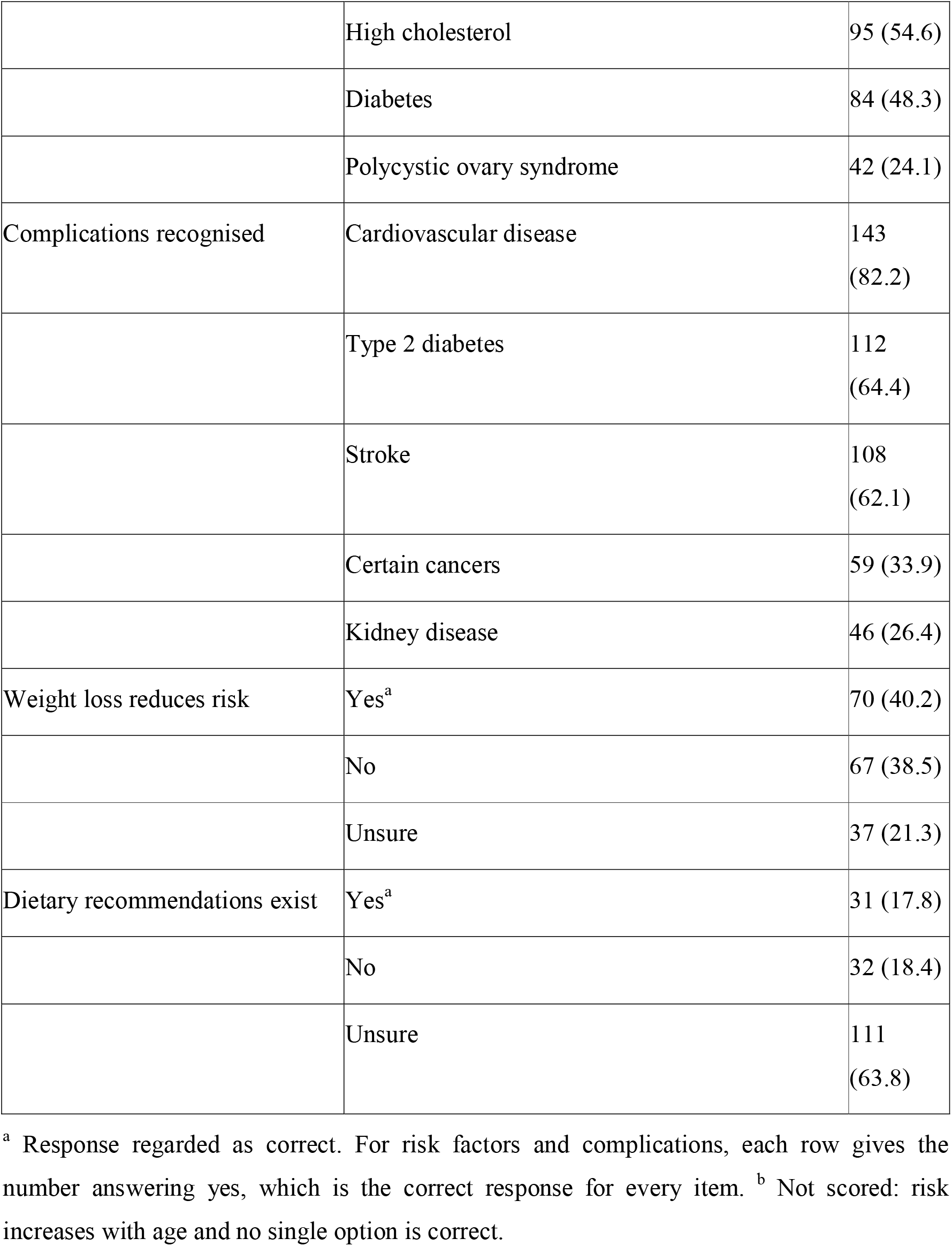
Knowledge of metabolic syndrome, its risk factors and complications (N = 174)

Recognition of individual risk factors varied widely. Poor diet (117; 67.2%), high blood pressure (116; 66.7%), obesity (114; 65.5%) and sedentary lifestyle (111; 63.8%) were recognised by about two-thirds of women, but smoking (49; 28.2%) and polycystic ovary syndrome (42; 24.1%) by fewer than one-third. Cardiovascular disease appeared to be the most widely recognised complication (143; 82.2%), followed by type 2 diabetes (112; 64.4%) and stroke (108; 62.1%); kidney disease (46; 26.4%) and certain cancers (59; 33.9%) appeared to be seldom recognised. Seventy women (40.2%) agreed that weight loss reduces the risk, and 111 (63.8%) were unsure whether dietary recommendations exist (Figure 1).

**Figure 1.**
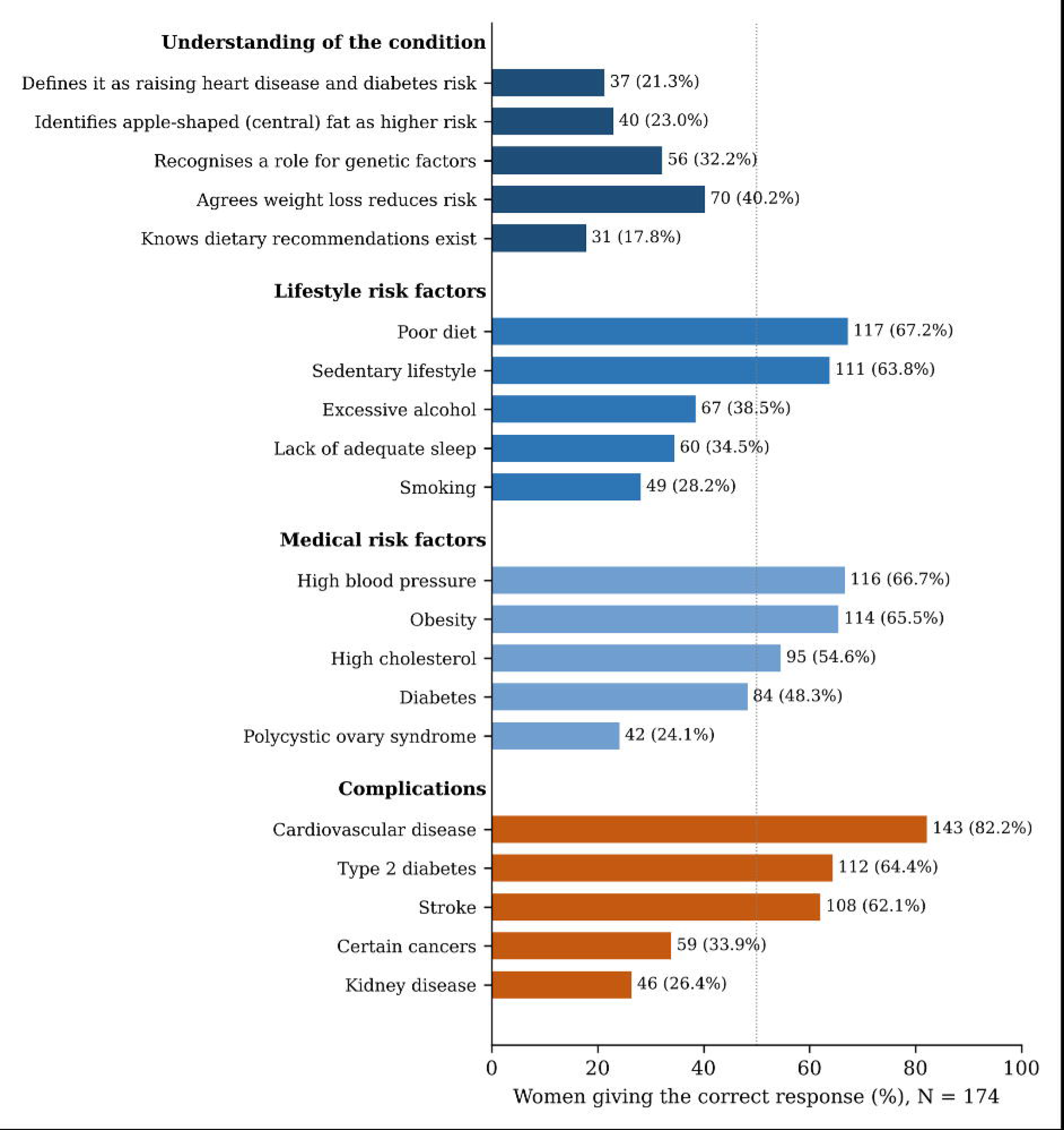
Proportion of women giving the correct response to each scored knowledge item on metabolic syndrome, its risk factors, and its complications (N = 174). Labels give the number and percentage of all respondents.

### 3.3 Lifestyle risk behaviours and self-reported conditions

Most women reported no weekly physical activity (116; 66.7%) and sedentary work (119; 68.4%). Daily consumption of sugary drinks (109; 62.6%) and processed or fast foods (100; 57.5%) appeared prevalent, while more than half of respondents rated their stress as high (103; 59.2%) and reported sleeping less than six hours a night (94; 54.0%). Most coped with stress through emotional support from family or friends (141; 81.0%), and only 9 (5.2%) sought counselling. Six women (3.4%) smoked regularly. Hypertension was the most frequently reported diagnosis (60; 34.5%), followed by type 2 diabetes (49; 28.2%), obesity (21; 12.1%), and high cholesterol (15; 8.6%) (Table 3).

**Table 3.** Lifestyle risk behaviours and self-reported diagnoses (N = 174)

| Item | Response | n (%) |
| --- | --- | --- |
| Physical activity per week | None | 116 (66.7) |
|  | Occasionally | 36 (20.7) |
|  | 3–4 times | 22 (12.6) |
| Type of physical activity (n = 138)<sup>a</sup> | Moderate | 83 (60.1) |
|  | Vigorous | 55 (39.9) |
| Sedentary work | Yes | 119 (68.4) |
| Sugary drinks | Daily | 109 (62.6) |
|  | 2–3 times a week | 30 (17.2) |
|  | Rarely | 35 (20.1) |
| Processed or fast foods | Daily | 100 (57.5) |
|  | 2–3 times a week | 33 (19.0) |
|  | Rarely | 41 (23.6) |
| Self-rated stress | High | 103 (59.2) |
|  | Moderate | 42 (24.1) |
|  | Low | 29 (16.7) |
| Main way of coping with stress | Support from family or friends | 141 (81.0) |
|  | Listening to music | 24 (13.8) |
|  | Seeking counselling | 9 (5.2) |
| Smoking | Yes, regularly | 6 (3.4) |
| Sleep per night | Less than 6 hours | 94 (54.0) |
|  | 6–7 hours | 26 (14.9) |
|  | 7–8 hours | 54 (31.0) |
| Ever diagnosed with | High blood pressure | 60 (34.5) |
|  | Type 2 diabetes | 49 (28.2) |
|  | Obesity | 21 (12.1) |
|  | High cholesterol | 15 (8.6) |

<sup>a</sup> As reported in the source analysis, this total is inconsistent with the 116 women who reported no physical activity (see Discussion). Percentages for this item use 138 as the denominator.

### 3.4 Preventive practices

Sixty-eight women (39.1%) reported recommending exercise three times a week for people with metabolic syndrome, whereas 60 (34.5%) recommended daily exercise. Fifty-nine (33.9%) had never received advice on prevention from a health professional, 101 (58.0%) had received it occasionally, and 14 (8.0%) regularly. Only 24 (13.8%) always followed a balanced diet, and 129 (74.1%) did so sometimes. Eighty-three (47.7%) undertook stress-reducing activities three to four times a week, and 25 (14.4%) rarely or never. Blood pressure was checked monthly by 53 (30.5%), every six months by 80 (46.0%), and rarely or never by 21 (12.1%). Most women (105, 60.3%) did not monitor their waist circumference or weight, and 167 (96.0%) had never been prescribed medication or supplements for metabolic risk (Table 4).

**Table 4.** Preventive knowledge and practices (N = 174)

| Item | Response | n (%) |
| --- | --- | --- |
| Recommended exercise frequency for people with metabolic syndrome | Daily | 60<br>(34.5) |
|  | 3 times per week | 68<br>(39.1) |
|  | 2 times per week | 46<br>(26.4) |
| Advice on prevention from a health professional | Yes, regularly | 14 (8.0) |
|  | Yes, occasionally | 101<br>(58.0) |
|  | Never | 59<br>(33.9) |
| Follows a balanced diet | Always | 24<br>(13.8) |
|  | Sometimes | 129<br>(74.1) |
|  | Rarely | 21<br>(12.1) |
| Stress-reducing activities | 3–4 times per week | 83<br>(47.7) |
|  | 1–2 times per week | 66<br>(37.9) |
|  | Rarely or never | 25<br>(14.4) |
| Prescribed medication or supplements | Yes, taken occasionally | 7 (4.0) |
|  | Never prescribed | 167<br>(96.0) |
| Blood pressure checked | Monthly | 53<br>(30.5) |
|  | Every 6 months | 80<br>(46.0) |
|  | Annually | 20<br>(11.5) |
|  | Rarely or never | 21<br>(12.1) |
| Monitors waist circumference and weight | Yes, regularly, and maintains healthy weight | 16 (9.2) |
|  | Yes, but does not always maintain healthy weight | 53<br>(30.5) |
|  | Does not monitor | 105<br>(60.3) |

Percentages may not sum to 100 because of rounding.

### 3.5 Lifestyle factors associated with self-reported diabetes and hypertension

Self-reported type 2 diabetes appeared to be associated with sleep duration (χ² = 6.70, df = 2, p = 0.035). It was reported by 36.2% of women sleeping less than 6 hours, compared with 15.4% of those sleeping 6–7 hours and 20.4% of those sleeping 7–8 hours. Diabetes also appeared to be associated with stress level (χ² = 7.17, df = 2, p = 0.028), but the proportion was highest among women reporting low stress (48.3%), compared with 21.4% with moderate and 25.2% with high stress. No association was observed between diabetes and physical activity (p = 0.665) or processed food consumption (p = 0.946) (Table 5; Figure 2).

**Figure 2.**
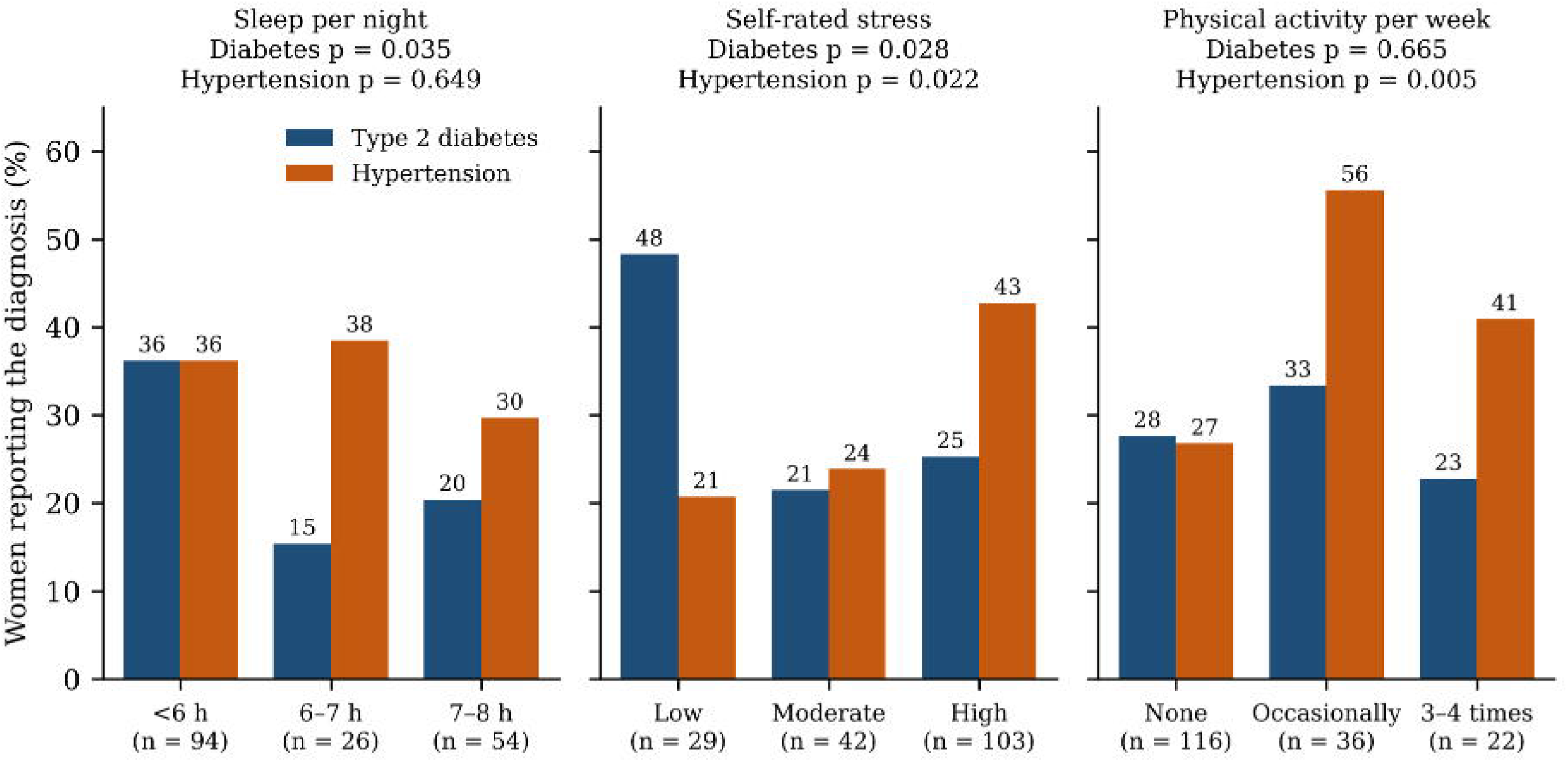
Proportion of women reporting a diagnosis of type 2 diabetes and of hypertension within categories of sleep duration, self-rated stress and weekly physical activity (N = 174). P-values are from Pearson’s chi-square tests (df = 2); associations are unadjusted.

**Table 5.** Self-reported type 2 diabetes and hypertension by lifestyle factor (N = 174)

| Lifestyle factor | Category | Women<br>category | Type 2 diabetes, n<br>(%) | Hypertension, n<br>(%) |
| --- | --- | --- | --- | --- |
| Physical activity per<br>week | None | 116 | 32 (27.6) | 31 (26.7) |
|  | Occasionally | 36 | 12 (33.3) | 20 (55.6) |
|  | 3–4 times | 22 | 5 (22.7) | 9 (40.9) |
| | $\chi^2$ (df = 2); p | | 0.82; 0.665 | 10.57; 0.005 |
| Self-rated stress | Low | 29 | 14 (48.3) | 6 (20.7) |
|  | Moderate | 42 | 9 (21.4) | 10 (23.8) |
|  | High | 103 | 26 (25.2) | 44 (42.7) |
| | $\chi^2$ (df = 2); p | | 7.17; 0.028 | 7.65; 0.022 |
| Sleep per night | Less than 6<br>hours | 94 | 34 (36.2) | 34 (36.2) |
|  | 6–7 hours | 26 | 4 (15.4) | 10 (38.5) |
|  | 7–8 hours | 54 | 11 (20.4) | 16 (29.6) |
| | $\chi^2$ (df = 2); p | | 6.70; 0.035 | 0.86; 0.649 |
| Processed or fast foods | Daily | 100 | 28 (28.0) | 33 (33.0) |
|  | 2–3 times a week | 33 | 10 (30.3) | 9 (27.3) |
|  | Rarely | 41 | 11 (26.8) | 18 (43.9) |
| | $\chi^2$ (df = 2); p | | 0.11; 0.946 | 2.47; 0.291 |
| All women |  | 174 | 49 (28.2) | 60 (34.5) |

Percentages are row percentages (women with the condition within each category). Pearson's chi-square tests, recomputed from the cross-tabulations reported in the source analysis; all expected counts were 5 or more. Associations are unadjusted.

Self-reported hypertension appeared to be associated with stress level (χ² = 7.65, df = 2, p = 0.022), being reported by 42.7% of women with high stress, compared with 23.8% with moderate and 20.7% with low stress. It also appeared to be associated with physical activity (χ² = 10.57, df = 2, p = 0.005), but in the opposite direction to that expected: hypertension was reported by 26.7% of women who did no physical activity, compared with 55.6% of those who were occasionally active and 40.9% of those active 3–4 times a week. No association was observed between hypertension and sleep duration (p = 0.649) or processed food consumption (p = 0.291).

## 4. Discussion

This survey of women attending a peri-urban polyclinic in Ghana found that few understood what metabolic syndrome is, while most reported several behaviours linked to it: physical inactivity, sedentary work, daily sugary drinks and fast food, high stress and short sleep. A third reported a diagnosis of hypertension and more than a quarter a diagnosis of diabetes. A third had never been advised about prevention, and most did not monitor their weight. Short sleep appeared to be associated with self-reported diabetes and high stress with self-reported hypertension, while two associations appeared to run in unexpected directions.

Only about one woman in five could describe metabolic syndrome correctly, and the most common answer confused it with a weight-loss diet. Fewer than a quarter recognised central fat as the high-risk pattern, a gap that may be of practical importance, since waist circumference appears to be the component women can most easily monitor themselves. The pattern of recognition suggests that women’s knowledge was built on familiar messages about diet, weight and blood pressure, with much less awareness of the condition as a whole or of less publicised links such as polycystic ovary syndrome, kidney disease and cancer. Uncertainty about dietary recommendations was widespread. Under the Health Belief Model, women who do not recognise a condition or the behaviours that raise their risk may be less likely to perceive themselves as susceptible, which may limit their motivation for preventive action (13).

The lifestyle profile may warrant attention in a group whose mean age was 35 years. Two-thirds reported no weekly physical activity, well short of the WHO recommendation of at least 150 minutes of moderate activity a week (11), and a similar proportion did sedentary work. Daily sugary drinks were reported by almost two-thirds; regular intake of sugar-sweetened beverages is associated with higher risks of metabolic syndrome and type 2 diabetes (8). These behaviours may be consistent with the dietary and occupational changes that accompany rapid urban growth in towns such as Kasoa. One inconsistency should be noted: 138 women reported a type of physical activity although 116 said they did none, which suggests that the activity-type question was answered by women who were not regularly active or was misunderstood.

The proportions reporting a diagnosis of hypertension (34.5%) and diabetes (28.2%) appear high for women aged 18–45 years. The diabetes figure in particular appears well above national estimates for Ghanaian adults (7). Women attending a health facility are more likely than the general population to have chronic conditions, and some may have been attending specifically for diabetes or hypertension care. Self-report may also be inaccurate in either direction: some women may have reported raised readings or gestational diabetes as diagnoses, and others may have undiagnosed conditions. These figures describe the clinic population and should not be read as community prevalence.

The association between short sleep and self-reported diabetes appears consistent with prospective evidence that sleeping less than six to seven hours is associated with a higher risk of type 2 diabetes (9). The association between high stress and self-reported hypertension is consistent with meta-analytic evidence linking psychosocial stress to hypertension (10). Both findings are unadjusted for age, education or other factors, and in a cross-sectional survey the direction cannot be established; living with a chronic condition can itself disturb sleep and raise stress.

Two associations ran against expectation. Women with self-reported diabetes more often rated their stress as low, and women with self-reported hypertension were more often physically active. A plausible explanation for the second is reverse causation: women already diagnosed with hypertension are commonly advised to exercise and may have increased their activity after diagnosis. A similar explanation may apply to stress if women with established diabetes had adapted to their condition, but this cannot be tested with the available data. These results may illustrate why cross-sectional associations between current behaviour and existing diagnoses cannot be read as risk factors for developing disease, and they should not be presented as such.

Preventive practice showed clear gaps. A third of women had never received advice on prevention from a health professional, although all were attending a health facility, and three in five did not monitor their weight or waist circumference. Most had their blood pressure checked at least every six months, which offers regular contact through which counselling could be given. Few women always followed a balanced diet, consistent with the high intake of sugary drinks and processed food.

This study adds evidence from a peri-urban primary-level facility in Ghana, where data on women’s understanding of metabolic syndrome are scarce (22). The findings suggest that knowledge gaps, a high prevalence of modifiable risk behaviours, and limited preventive counselling coexist among women of reproductive age who are already in contact with health services. The findings may be relevant to nurses, midwives, and managers at Kasoa Polyclinic and to the Awutu Senya East Municipal Health Directorate in planning health education and screening.

## 5. Strengths and limitations

The study covered knowledge, behaviour, self-reported diagnoses and preventive practice in one sample of women, used a pre-tested questionnaire administered with language support so that women without formal education could take part, and achieved its calculated sample size.

Several limitations affect interpretation. Convenience sampling at one facility means the women may differ from those in the wider Kasoa population, and the response rate is unknown. Metabolic syndrome was not measured: no waist circumference, blood pressure or laboratory tests were taken, so the study describes knowledge and risk behaviours and cannot estimate the prevalence of the syndrome. Diagnoses and behaviours were self-reported and subject to recall and social desirability bias. The knowledge items had not been validated, and the analysis could only be reported item by item. The associations are bivariate and unadjusted, so confounding by age, education, occupation or the reason for attendance is likely; the cross-sectional design prevents conclusions about the direction of any association. Several categories of the original questions received no responses or were combined, and the questions on physical activity frequency and type gave inconsistent totals.

## 6. Implications for practice, policy and research

For practice, the polyclinic’s regular contact with women for blood pressure checks and other services offers an opportunity for brief structured counselling on metabolic risk. This could explain what metabolic syndrome is, emphasise central obesity and show women how to measure their waist. It should also address sugary drinks, processed foods, physical activity and sleep. Nurses and midwives could integrate waist and weight measurement into routine visits for women of reproductive age.

For programmes, the Municipal Health Directorate could include metabolic syndrome messages in existing non-communicable disease and reproductive health education, with attention to the gap in knowledge about central fat distribution.

For research, community-based studies in Kasoa should measure the components of metabolic syndrome directly, using probability sampling and validated knowledge scales. Longitudinal designs may be needed to establish whether short sleep and stress precede the onset of diabetes and hypertension in this population (23), and analyses should adjust for age and socio-economic factors.

## 7. Conclusion

Among women aged 18–45 years attending Kasoa Polyclinic, understanding of metabolic syndrome was limited, particularly regarding the nature of the condition and the importance of central obesity. Physical inactivity, sedentary work, daily sugary drinks and fast food, high stress and short sleep were common, and a third of women had never received preventive advice. Self-reported diabetes was associated with short sleep and self-reported hypertension with high stress, but these unadjusted associations need confirmation with measured outcomes. Structured counselling and routine waist and weight monitoring at the polyclinic are reasonable first steps.

## Declarations

### Ethical approval

The study was approved by the Ghana Health Service Ethics Review Committee (GHS-ERC: 047/04/24; approval date 25 June 2024). Permission was granted by the management of Kasoa Polyclinic and the Awutu Senya East Municipal Health Directorate.

### Consent

Written informed consent, by signature or thumbprint, was obtained from all participants before data collection.

### Authors’ contributions

Grace De-Graft Aitee led the conceptualization, methodology, statistical analysis, and preparation of the original draft; Abena Ntiriwaa Cann and Edmund Teye Borkor contributed to data curation and validation; Abubakar Iddrisu Siddiq contributed to the literature review and interpretation of findings; Afua Pokuaa Kwakye contributed to data visualization and results interpretation; Ernest Alfred Kwaku Ampong contributed to methodological review and critical revision of the manuscript; and Evelyn Owusu contributed to supervision, project administration, and final manuscript review, with all six authors approving the final version.

## Data availability

The data that support the findings are available from the corresponding author on reasonable request, subject to the conditions of the ethical approval.

## Acknowledgements

The authors thank the women who took part, the management and staff of Kasoa Polyclinic, the research assistants, and Mrs Martha Atiemo, Faculty Chair of Well Woman Care, Ghana College of Nurses and Midwives, for her advice.

## References

1. Alberti KGMM, Eckel RH, Grundy SM, Zimmet P, Cleeman JI, Donato KA, et al. Harmonizing the Metabolic Syndrome. Circulation. 2009 Oct 5;120(16):1640–5. doi: 10.1161/circulationaha.109.192644 PubMed PMID: 19805654.

2. Naranjo-Díaz MB, Borbor-Soriano JA, Bazán-Magallán WA, Panchana-Vera DV. Influencia del sedentarismo y la dieta occidental en el desarrollo del síndrome metabólico: una revisión de literatura. MQRInvestigar. 2024 Nov 30;8(4):5474–503. doi: 10.56048/mqr20225.8.4.2024.5474-5503

3. Noubiap JJ, Nansseu JR, Lontchi Yimagou E, Nkeck JR, Nyaga UF, Ngouo AT, et al. Geographic distribution of metabolic syndrome and its components in the general adult population: A meta-analysis of global data from 28 million individuals. Diabetes Research and Clinical Practice. 2022 May 15;188:109924. doi: 10.1016/j.diabres.2022.109924 PubMed PMID: 35584716.

4. Bowo Ngandji A, Kenmoe S, Ebogo Belobo JT, Kenfack Momo R, Takuissu GR, Kengne-Ndé C, et al. Prevalence of the metabolic syndrome in African populations: A systematic review and meta-analysis. PLoS ONE. 2023 Jul 27;18(7). doi: 10.1371/journal.pone.0289155 PubMed PMID: 37498832.

5. Ofori Asenso R, Agyeman AA, Laar AK. Metabolic Syndrome in Apparently “Healthy” Ghanaian Adults: A Systematic Review and Meta-Analysis. International Journal of Chronic Diseases. 2017 Jan 1;2017:1–9. doi: 10.1155/2017/2562374 PubMed PMID: 29130065.

6. Atibila F, Hoor GA ten, Donkoh ET, Abdul IW, Kok G. Prevalence of hypertension in Ghanaian society: a systematic review, meta-analysis, and GRADE assessment. Systematic Reviews. 2021 Aug 7;10(1):220. doi: 10.1186/s13643-021-01770-x PubMed PMID: 34364395.

7. Asamoah-Boaheng M, Sarfo Kantanka O, Tuffour AB, Eghan B, Mbanya JC. Prevalence and risk factors for diabetes mellitus among adults in Ghana: a systematic review and meta-analysis. International Health. 2018 Sep 19;11(2):83–92. doi: 10.1093/inthealth/ihy067 PubMed PMID: 30285118.

8. Malik V, Popkin BM, Bray GA, Després J, Willett WC, Hu FB. Sugar-Sweetened Beverages and Risk of Metabolic Syndrome and Type 2 Diabetes. Diabetes Care. 2010 Aug 6;33(11):2477–83. doi: 10.2337/dc10-1079 PubMed PMID: 20693348.

9. Shan Z, Ma H, Xie M, Yan P, Guo Y, Bao W, et al. Sleep Duration and Risk of Type 2 Diabetes: A Meta-analysis of Prospective Studies. Diabetes Care. 2015 Feb 12;38(3):529–37. doi: 10.2337/dc14-2073 PubMed PMID: 25715415.

10. Liu M, Li N, Li WA, Khan H. Association between psychosocial stress and hypertension: a systematic review and meta-analysis. Neurological Research. 2017 Apr 17;39(6):573–80. doi: 10.1080/01616412.2017.1317904 PubMed PMID: 28415916.

11. Bull F, Al-Ansari SS, Biddle S, Borodulin K, Buman MP, Cardon G, et al. World Health Organization 2020 guidelines on physical activity and sedentary behaviour. British Journal of Sports Medicine. 2020 Nov 25;54(24):1451–62. doi: 10.1136/bjsports-2020-102955 PubMed PMID: 33239350.

12. Yussif MT, Morrison AE, Annan RA. 10-year level, trends and socio-demographic disparities of obesity among Ghanaian adults—A systematic review and meta-analysis of observational studies. PLOS Global Public Health. 2024 Jan 25;4(1). doi: 10.1371/journal.pgph.0002844 PubMed PMID: 38271466.

13. Champion VL, Skinner CS. The health belief model. [Internet]. 2008 Jan 1 [cited 2026 Sep]. Available from: https://psycnet.apa.org/record/2008-17146-003

14. Kazibwe J, Gad M, Abassah-Konadu E, Amankwah I, Owusu R, Gulbi G, et al. The epidemiological and economic burden of diabetes in Ghana: A scoping review to inform health technology assessment. PLOS Global Public Health. 2024 Mar 12;4(3). doi: 10.1371/journal.pgph.0001904 PubMed PMID: 38470940.

15. Poku AA, Sarkodie K, Mensah J, Sam-Mensah R, Quayson JJ, Poku K. Livelihood resilience among street hawkers during Covid-19 lockdown in the Awutu Senya East Municipality, Ghana. Journal of Humanities and Applied Social Sciences. 2024 Jun 10;7(3):220–35. doi: 10.1108/jhass-09-2023-0125

16. Elm EB von, Altman DG, Egger M, Pocock S, Gøtzsche PC, Vandenbroucke JP. The Strengthening the Reporting of Observational Studies in Epidemiology (STROBE) statement: guidelines for reporting observational studies. The Lancet. 2007 Oct 1;370(9596):1453–7. doi: 10.1016/s0140-6736(07)61602-x PubMed PMID: 18064739.

17. Cochran WG. Sampling Techniques. 3rd Edition. Zenodo (CERN European Organization for Nuclear Research). 1977 Jan 1. doi: 10.5281/zenodo.15338807

18. Gazzaz ZJ. Knowledge, Attitudes, and Practices Regarding Diabetes Mellitus Among University Students in Jeddah, Saudi Arabia. Diabetes Metabolic Syndrome and Obesity. 2020 Dec 1;5071–8. doi: 10.2147/dmso.s287459 PubMed PMID: 33380816.

19. Agbozo F, Bannerman E, Klomegah S, Zotor F. Lifestyle habits and perceived wellbeing of adults presenting with metabolic syndrome at a diabetic clinic in Ghana: A case-control study. Human Nutrition & Metabolism. 2022 Jun 8;29:200154. doi: 10.1016/j.hnm.2022.200154

20. Boakye H, Atabila A, Hinneh T, Ackah M, Ojo-Benys F, Bello A. The prevalence and determinants of non-communicable diseases among Ghanaian adults: A survey at a secondary healthcare level. PLoS ONE. 2023 Feb 8;18(2). doi: 10.1371/journal.pone.0281310 PubMed PMID: 36753484.

21. Bae C, Piao M, Kim MY, Im Y, Kim S, Kim DU, et al. Biological age and lifestyle in the diagnosis of metabolic syndrome: the NHIS health screening data, 2014–2015. Scientific Reports. 2021 Jan 11;11(1):444. doi: 10.1038/s41598-020-79256-4 PubMed PMID: 33431923.

22. Achempim-Ansong G, Tshabalala AM, Gradidge PJL. Factors Associated with Improved Knowledge of Metabolic Syndrome in Female Market Traders. International Journal of Environmental Research and Public Health. 2022 Sep 27;19(19):12256. doi: 10.3390/ijerph191912256 PubMed PMID: 36231550.

23. Kivimäki M, Bartolomucci A, Kawachi I. The multiple roles of life stress in metabolic disorders. PubMed Central. 2022 Oct 12;19(1):10–27. doi: 10.1038/s41574-022-00746-8 PubMed PMID: 36224493.

